# Predicting Infant Nonattendance at the Next Recommended Well-Child Visit: Model Development and Validation

**DOI:** 10.64898/2026.03.24.26348063

**Authors:** Amanda Luff, Maureen Shields, Jana Hirschtick, Marybeth Ingle, Clare Crosh, Melanie Marsh, François Modave, Veronica Fitzpatrick

## Abstract

**Background:** Well-child visits (WCVs) are essential for monitoring infant development, yet nonattendance remains common. Machine learning models have predicted appointment no-shows, but these approaches require a visit to be scheduled and cannot identify children whose visits are never scheduled or are cancelled in advance. We developed and validated models that predict infant nonattendance at the next recommended WCV using information available at the current visit.

**Methods:** We used visit-level electronic health record data from two demographically distinct pediatric practices: Practice A (5,300 visits, 1,215 patients) served as the training and internal validation set; Practice B (5,274 visits, 1,271 patients) served as the external validation set. We compared regularized logistic regression, random forest, and XGBoost using group k-fold cross-validation. Model discrimination was assessed using AUC and average precision, and threshold-dependent metrics were reported at the F1-maximizing threshold. We evaluated subgroup performance by race/ethnicity, preferred language, and insurance type in the external validation set.

**Results:** All three models achieved similar discrimination in internal validation (AUC 0.69-0.72, average precision 0.36-0.41) and external validation (AUC 0.66-0.68, average precision 0.36-0.38). Across all models, care delivery features, including visit timepoint, visit delay, scheduling lead time, and prior no-shows, were the most influential predictors. Logistic regression with LASSO regularization retained six nonzero coefficients, suggesting comparable performance is achievable with a reduced feature set. Model discrimination was similar across insurance groups, between English– and Spanish-speaking patients, and between Hispanic and non-Hispanic Black patients relative to non-Hispanic white patients. Estimates for smaller subgroups were imprecise due to limited sample sizes.

**Conclusions:** Predicting WCV nonattendance at the next recommended timepoint is feasible using routine visit data. A parsimonious logistic regression model performed comparably to more complex algorithms. Because prediction occurs during an active visit, clinicians can address barriers to the next visit while the family is still present.

## Background

Well-child visits (WCVs) are essential for monitoring pediatric development and health and create early connections between clinicians and families.^1^ The American Academy of Pediatrics (AAP) recommends all children receive a series of preventative visits at prespecified timepoints in their first year of life. Missed WCVs have been associated with increased rates of preventable acute care visits, delayed recognition of developmental delays, and missed opportunities for early intervention.^2–4^ Although WCVs are important for ongoing pediatric health, there is variability in visit attendance by sociodemographic and health status factors.^5–7^ Patients who identify as Hispanic or non-Hispanic Black, have public insurance, and live in lower income areas are more likely to miss WCVs.

Although the importance of WCVs is widely known, barriers to attendance persist, including financial stress, work constraints, childcare, transportation, and language barriers. ^8–10^ Additionally, health system and provider-level constraints exist, such as appointment availability and insufficient time during visits to cover care needs.^11^ Reminder systems, such as phone calls, text messages, and patient portal messages, are common strategies used to improve visit attendance among health systems. However, these methods tend to be resource-intensive and are often implemented uniformly rather than tailored to individual family risk.^12–14^ As a consequence, improvements in attendance are often modest, and disparities in preventive care utilization remain.^7,14^

Machine learning (ML) has been increasingly used in healthcare in the form of clinical decision support tools and risk stratification models, which use patient-specific data to inform care.^15,16^ ML models have been applied to predict appointment no-shows across clinical settings, including pediatrics, integrating demographic, behavioral, and clinical data to identify patients at risk of missing scheduled visits.^17–20^ However, no-show models address the narrower question of whether a patient with a scheduled appointment will attend that specific appointment. This approach conditions on a visit being scheduled and cannot capture children whose visits are never scheduled or cancelled in advance.

Most prior ML research in pediatrics has focused on acute care outcomes or disease prediction rather than preventive care delivery.^21–23^ There is limited evidence on the use of ML to predict WCV nonattendance, as well as insufficient understanding of how such tools perform across the diverse patient populations they are intended to serve.^22,23^ We aimed to develop and validate ML models to predict whether a child will complete a well-child visit at the next AAP-recommended timepoint, using information available at the current visit. Unlike no-show prediction, this outcome is agnostic to whether the next visit was ever scheduled, cancelled, or no-showed, and instead captures whether the child received timely preventive care at the recommended interval. We trained models on one practice and validated on a second, demographically distinct practice, and assessed model discrimination across racial/ethnic, linguistic, and insurance subgroups. An effective predictive tool could enable pediatricians to identify potential barriers to care and offer targeted support in real time, while the family is still present and engaged in the medical home.

## Methods

We assessed WCV attendance at pediatric practices affiliated with two Chicago-area children’s hospitals. Practice A, used for model training and internal validation, serves patients in the north side of the Chicago area. The patient population is predominantly white and Asian, and treating providers are physician attendings and residents. Practice B, used for external validation, serves patients in the south side of the Chicago area. The patient population is predominantly Black and Hispanic, with most patients having Medicaid insurance, and treating providers include physician attendings, residents, and Advance Practice Providers (APP). Further information about this cohort has been previously published.^6,24^ This analysis uses visit-level data from the included patients, with each patient contributing 1-7 visits to the data set.

The Advocate Health Wake Forest University School of Medicine Institutional Review Board approved this research and waived the informed consent requirement due to the retrospective methods.

### Outcome

We identified WCVs using the SNOMED CT concept for a WCV. SNOMED is a standardized set of EHR clinical concepts used to represent encounter context (i.e., the clinical purpose of the visit) and enables consistent recording and sharing of detailed clinical data. While prior retrospective research often uses ICD-10 codes and CPT codes to identify WCVs, these codes are primarily used for billing and reporting and may be administratively modified for billing compliance after the encounter. On the other hand, SNOMED codes capture clinical context, which is known by the clinician at point of care and better mirrors data that would be available to clinicians in real-world implementation. SNOMED codes are able to capture greater granularity than ICD-10 codes, with SNOMED containing over 350,000 clinical concepts, while ICD-10 contains fewer than 100,000.^25^ Further, codes primarily used for billing, such as CPT, are often poorly populated in EHR-derived datasets.^26^

The AAP recommended timepoints for infant WCVs are newborn, 1, 2, 4, 6, 9, 12, and 15 months. Visits through 12 months were included in this analysis, with attendance at the 15-month visit serving as the outcome for the 12-month visit. Because there are no standardized windows for WCVs, assigning visits to specific timepoints may differ between studies. Here, we define a missed next visit as one not completed before the subsequent recommended timepoint. For example, if no visit occurs between 6 and under 9 months, the 6-month visit is missed.

### Data Preprocessing and Feature Creation

Predictors included visit-level features (age at visit, scheduling lead time, first visit at the clinic, provider type), prior utilization (previous missed WCV, any no-show within the health system, emergency department visit, documented immunization refusal), and patient characteristics (sex, insurance type, preferred language). Where available, we assessed birth record information, including low birthweight (<2,500 grams), preterm birth (<37 weeks gestation), and NICU admission. Patients without a birth record in our system were coded as 0 for all birth variables, with a separate indicator used to distinguish true negatives from out-of-system births. Race and ethnicity are described but were not included as model features.

### Model Development

We compared three predictive ML models by their ability to accurately predict non-attendance: regularized logistic regression, random forests, and XGBoost.^27,28^ In previous analyses of this patient population, we observed missed-visit rates of 15-35%.^6^ To account for the lower proportion of subsequent missed compared to completed visits (i.e. class imbalance), we used Synthetic Minority Over-sampling TEchnique (SMOTE) for tree-based models^29^ and specified class weight as balanced for logistic regression models. We used nested group-k fold cross validation grouped by patient to avoid data leakage, which would occur if data from one patient was present in both the training and testing folds. Average Precision was used for model scoring because this metric prioritizes both recall and precision and is preferred over AUC for imbalanced data. Hyperparameter search strategies are displayed in **S1 Table**. Following initial model creation, we identified features to remove that did not contribute to the model’s predictive ability using a permutation importance threshold of 0 and dropping features with negative values.

To assess model performance, we generated ROC curves and Precision-Recall curves for out-of-fold predictions on Practice A (internal validation) and for Practice B (external validation). We additionally report sensitivity (recall), specificity, negative predictive value, positive predictive value (precision), accuracy, balanced accuracy, and F1 score for each of the internal and external validation sets. All threshold-dependent metrics are reported at the threshold that maximizes F1 score, a metric that balances precision and recall. We secondarily report threshold-specific metrics at a 0.5 threshold. 95% confidence intervals for all performance metrics were estimated using the percentile bootstrap (2,000 replicates). For threshold-dependent metrics, the classification threshold was determined on the full sample and held fixed across replicates.

To support interpretability, we calculated SHAP values and generated bee swarm plots for internal and external validation sets, explains which pieces of information pushed the model toward or away from predicting a missed visit for a given patient, and by how much.^30^ This allowed us to understand which factors were most important in predicting WCV attendance.

To assess whether model discrimination differed across patient subgroups, we computed AUC stratified by combined race and ethnicity, preferred language, and insurance type in the external validation set. Race and ethnicity were not included as model features. Preferred language was evaluated during initial model development but removed from all models after achieving negative permutation importance. We report the AUC gap, defined as the difference between the highest and lowest subgroup AUC within each grouping variable, as a summary measure of performance equity.

All analyses were completed using Python (Version 3.1; Python Software Foundation, 2016).

## Results

The training set (Practice A) included 1,215 unique patients (5,300 visits); the external validation set (Practice B) included 1,271 unique patients (5,274 visits). The proportion of missed next visits was 16.2% in Practice A and 20.7% in Practice B **(Table 1)**.

**TABLE 1.** Training and Testing Cohort Characteristics (Visit-level)

|  |  | Train (Practice A) |  | Test (Practice B) |  |
| --- | --- | --- | --- | --- | --- |
|  | Variable | N | % | N | % |
| **Next Visit is Missed (Outcome)** | No | 4439 | (83.8) | 4180 | (79.3) |
|  | Yes | 861 | (16.2) | 1094 | (20.7) |
| **Timepoint** | 0 months | 175 | (3.3) | 166 | (3.1) |
|  | 1 months | 646 | (12.2) | 702 | (13.3) |
|  | 2 months | 883 | (16.7) | 900 | (17.1) |
|  | 4 months | 885 | (16.7) | 893 | (16.9) |
|  | 6 months | 944 | (17.8) | 974 | (18.5) |
|  | 9 months | 818 | (15.4) | 784 | (14.9) |
|  | 12 months | 949 | (17.9) | 855 | (16.2) |
| **Prior Missed WCV in Dataset** | No | 4050 | (76.4) | 3953 | (75.0) |
|  | Yes | 1250 | (23.6) | 1321 | (25.0) |
| **Documented Immunization Refusal on or before date** | No | 5036 | (95.0) | 4994 | (94.7) |
|  | Yes | 264 | (5.0) | 280 | (5.3) |
| **ED Visit before date** | No | 4422 | (83.4) | 4372 | (82.9) |
|  | Yes | 878 | (16.6) | 902 | (17.1) |
| **No-Show before date** | No | 4408 | (83.2) | 4340 | (82.3) |
|  | Yes | 892 | (16.8) | 934 | (17.7) |
| **First Visit with Clinic** | No | 4969 | (93.8) | 4867 | (92.3) |
|  | Yes | 331 | (6.2) | 407 | (7.7) |
| **Patient Portal Active** | No | 1922 | (36.3) | 2200 | (41.7) |
|  | Yes | 3378 | (63.7) | 3074 | (58.3) |
| **Provider is Physician (Attending, Resident, Fellow)** | No | 936 | (17.7) | 0 | (0.0) |
|  | Yes | 4364 | (82.3) | 5274 | (100.0) |
| **Provider is Physician Attending** | No | 1347 | (25.4) | 1245 | (23.6) |
|  | Yes | 3953 | (74.6) | 4029 | (76.4) |
| **Sex** | Female | 2540 | (47.9) | 2555 | (48.4) |
|  | Male | 2760 | (52.1) | 2719 | (51.6) |
| **Insurance** | Not Medicaid | 2849 | (53.8) | 1469 | (27.9) |
|  | Medicaid | 2451 | (46.2) | 3805 | (72.1) |
| **Has Birth Record** | No | 1221 | (23.0) | 1054 | (20.0) |
|  | Yes | 4079 | (77.0) | 4220 | (80.0) |
| **Documented Low Birth Weight <sup>a</sup>** | No | 4908 | (92.6) | 4742 | (89.9) |
|  | Yes | 392 | (7.4) | 532 | (10.1) |
| **Documented NICU Admission <sup>a</sup>** | No | 4742 | (89.5) | 5268 | (99.9) |
|  | Yes | 558 | (10.5) | 6 | (0.1) |
| **Documented Preterm Birth <sup>a</sup>** | No | 4802 | (90.6) | 4566 | (86.6) |
|  | Yes | 498 | (9.4) | 708 | (13.4) |
| **Preferred Language <sup>b</sup>** | English | 5011 | (94.5) | 4913 | (93.2) |
|  | Spanish | 136 | (2.6) | 208 | (3.9) |
|  | Another Language | 153 | (2.9) | 153 | (2.9) |
| **Race <sup>c</sup>** | White | 3298 | (62.2) | 2432 | (46.1) |
|  | Black | 216 | (4.1) | 2465 | (46.7) |
|  | Asian | 1145 | (21.6) | 77 | (1.5) |
|  | Another Race | 475 | (9.0) | 201 | (3.8) |
|  | Not Reported | 166 | (3.1) | 99 | (1.9) |
| **Ethnicity <sup>c</sup>** | Hispanic or Latino | 1636 | (30.9) | 1969 | (37.3) |
|  | Not Hispanic or Latino | 3575 | (67.5) | 3240 | (61.4) |
|  | Not Reported | 89 | (1.7) | 65 | (1.2) |

<sup>a</sup> Only positive if patient present and if patient had a birth record
<sup>b</sup> Removed after initial model fit due to negative permutation importance value
<sup>c</sup> Not included in model training

After initial model training, preferred language was uninformative (e.g., achieved a negative permutation importance score for RF and XGBoost) and thus was removed from all models. Under the ideal hyperparameters following a grid search, the logistic regression model with L1 regularization (LASSO) retained six coefficients that meaningfully contributed to prediction of WCV nonattendance: timepoint, visit delay, prior no-show, schedule lead time in days, new to department, and prior immunization refusal.

In the internal validation set, the random forest and XGBoost models achieved marginally higher AUC and average precision than the logistic regression model, indicating that these models were slightly better at ordering patients from lower to higher risk across the full range of predicted scores (**Table 2**, **Figure 1**). In contrast, logistic regression achieved the highest maximized F1 score, meaning that when predictions were converted into a binary classification, it provided the most favorable balance between correctly identifying patients with the outcome (recall) and limiting incorrect positive classifications (precision).

**FIGURE 1.**
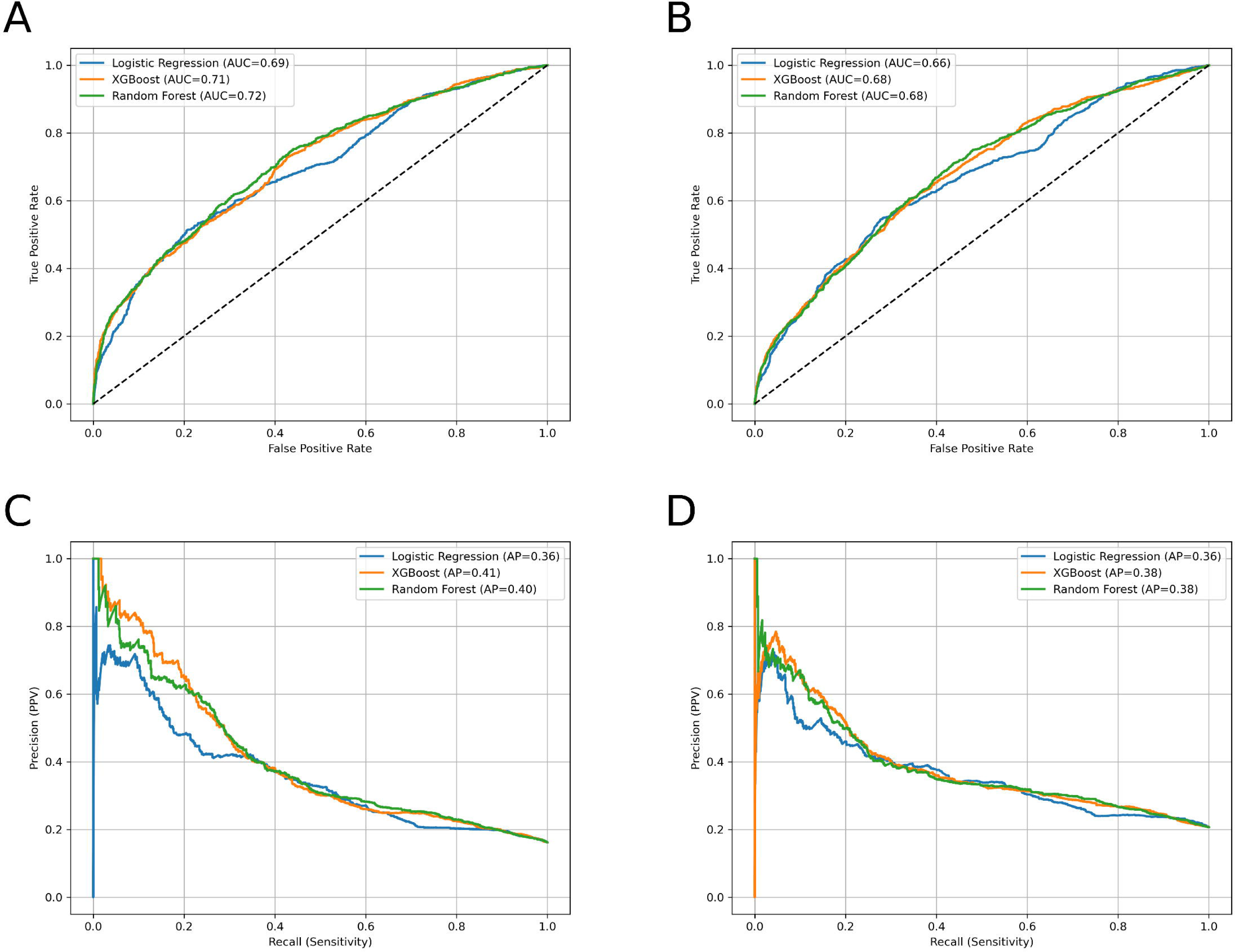
Diagnostic Plots for Internal and External Validation Sets. **Legend:** The top row shows receiver operating characteristic (ROC) curves (A, B), and the bottom row shows precision–recall curves (C, D). The left column presents internal validation using out of fold predictions from Practice A (A, C), and the right column presents external validation using Practice B (B, D).

**TABLE 2.**
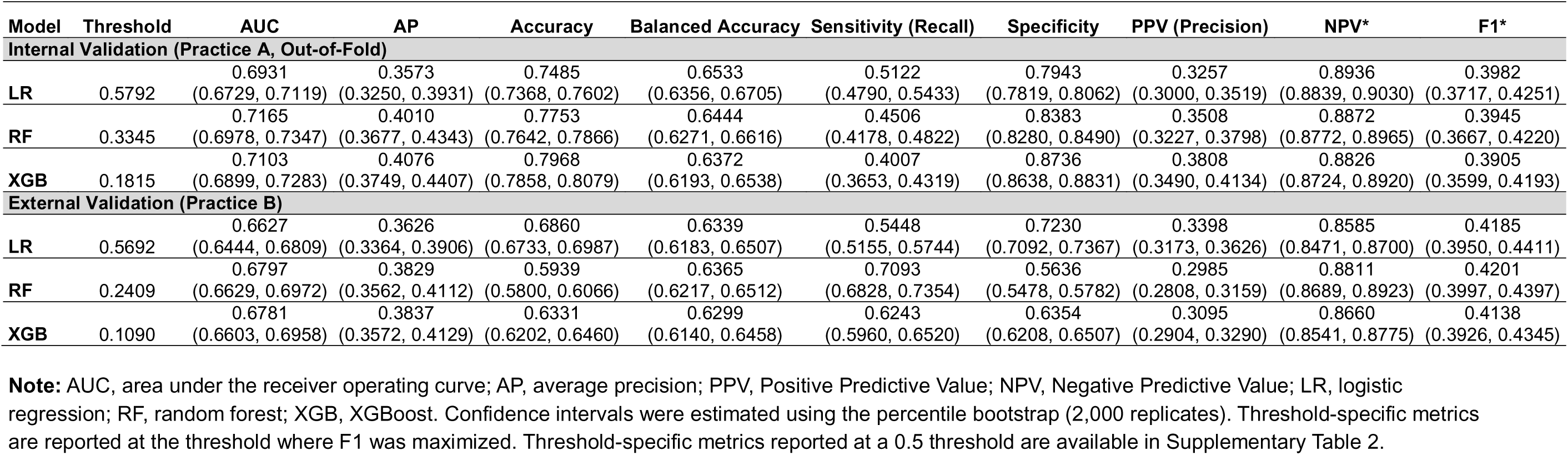
Model Diagnostics for Internal and External Validation Sets.

| Model | Threshold | AUC | AP | Accuracy | Balanced Accuracy | Sensitivity (Recall) | Specificity | PPV (Precision) | NPV* | F1* |
| --- | --- | --- | --- | --- | --- | --- | --- | --- | --- | --- |
| Internal Validation (Practice A, Out-of-Fold) |  |  |  |  |  |  |  |  |  |  |
| LR | 0.5792 | 0.6931 | 0.3573 | 0.7485 | 0.6533 | 0.5122 | 0.7943 | 0.3257 | 0.8936 | 0.3982 |
|  |  | (0.6729, 0.7119) | (0.3250, 0.3931) | (0.7368, 0.7602) | (0.6356, 0.6705) | (0.4790, 0.5433) | (0.7819, 0.8062) | (0.3000, 0.3519) | (0.8839, 0.9030) | (0.3717, 0.4251) |
| RF | 0.3345 | 0.7165 | 0.4010 | 0.7753 | 0.6444 | 0.4506 | 0.8383 | 0.3508 | 0.8872 | 0.3945 |
|  |  | (0.6978, 0.7347) | (0.3677, 0.4343) | (0.7642, 0.7866) | (0.6271, 0.6616) | (0.4178, 0.4822) | (0.8280, 0.8490) | (0.3227, 0.3798) | (0.8772, 0.8965) | (0.3667, 0.4220) |
| XGB | 0.1815 | 0.7103 | 0.4076 | 0.7968 | 0.6372 | 0.4007 | 0.8736 | 0.3808 | 0.8826 | 0.3905 |
|  |  | (0.6899, 0.7283) | (0.3749, 0.4407) | (0.7858, 0.8079) | (0.6193, 0.6538) | (0.3653, 0.4319) | (0.8638, 0.8831) | (0.3490, 0.4134) | (0.8724, 0.8920) | (0.3599, 0.4193) |
| External Validation (Practice B) |  |  |  |  |  |  |  |  |  |  |
| LR | 0.5692 | 0.6627 | 0.3626 | 0.6860 | 0.6339 | 0.5448 | 0.7230 | 0.3398 | 0.8585 | 0.4185 |
|  |  | (0.6444, 0.6809) | (0.3364, 0.3906) | (0.6733, 0.6987) | (0.6183, 0.6507) | (0.5155, 0.5744) | (0.7092, 0.7367) | (0.3173, 0.3626) | (0.8471, 0.8700) | (0.3950, 0.4411) |
| RF | 0.2409 | 0.6797 | 0.3829 | 0.5939 | 0.6365 | 0.7093 | 0.5636 | 0.2985 | 0.8811 | 0.4201 |
|  |  | (0.6629, 0.6972) | (0.3562, 0.4112) | (0.5800, 0.6066) | (0.6217, 0.6512) | (0.6828, 0.7354) | (0.5478, 0.5782) | (0.2808, 0.3159) | (0.8689, 0.8923) | (0.3997, 0.4397) |
| XGB | 0.1090 | 0.6781 | 0.3837 | 0.6331 | 0.6299 | 0.6243 | 0.6354 | 0.3095 | 0.8660 | 0.4138 |
|  |  | (0.6603, 0.6958) | (0.3572, 0.4129) | (0.6202, 0.6460) | (0.6140, 0.6458) | (0.5960, 0.6520) | (0.6208, 0.6507) | (0.2904, 0.3290) | (0.8541, 0.8775) | (0.3926, 0.4345) |

**Note:** AUC, area under the receiver operating curve; AP, average precision; PPV, Positive Predictive Value; NPV, Negative Predictive Value; LR, logistic regression; RF, random forest; XGB, XGBoost. Confidence intervals were estimated using the percentile bootstrap (2,000 replicates). Threshold-specific metrics are reported at the threshold where F1 was maximized. Threshold-specific metrics reported at a 0.5 threshold are available in Supplementary Table 2.

In the external validation set, random forest and XGBoost again showed higher AUC and average precision than logistic regression, but the smaller differences indicate that overall risk discrimination was more similar across models in the external population. Maximized F1 scores were also similar across models, with random forest performing slightly better than the others, indicating only modest differences in the balance of recall and precision.

At the threshold which maximized F1 score, recall ranged from 0.40-0.51 in the internal valuation set and 0.54-0.71 in the external validation set.

Using SHAP values, all models identified timepoint, visit delay, schedule lead days, and prior no-show as among the top 6 most influential features (**Figure 2**). In all models, visits at an earlier timepoint (younger age) and a larger visit delay (greater time between the recommended visit month and actual visit time), and prior no-shows were associated with prediction that a visit was missed.

**FIGURE 2.**
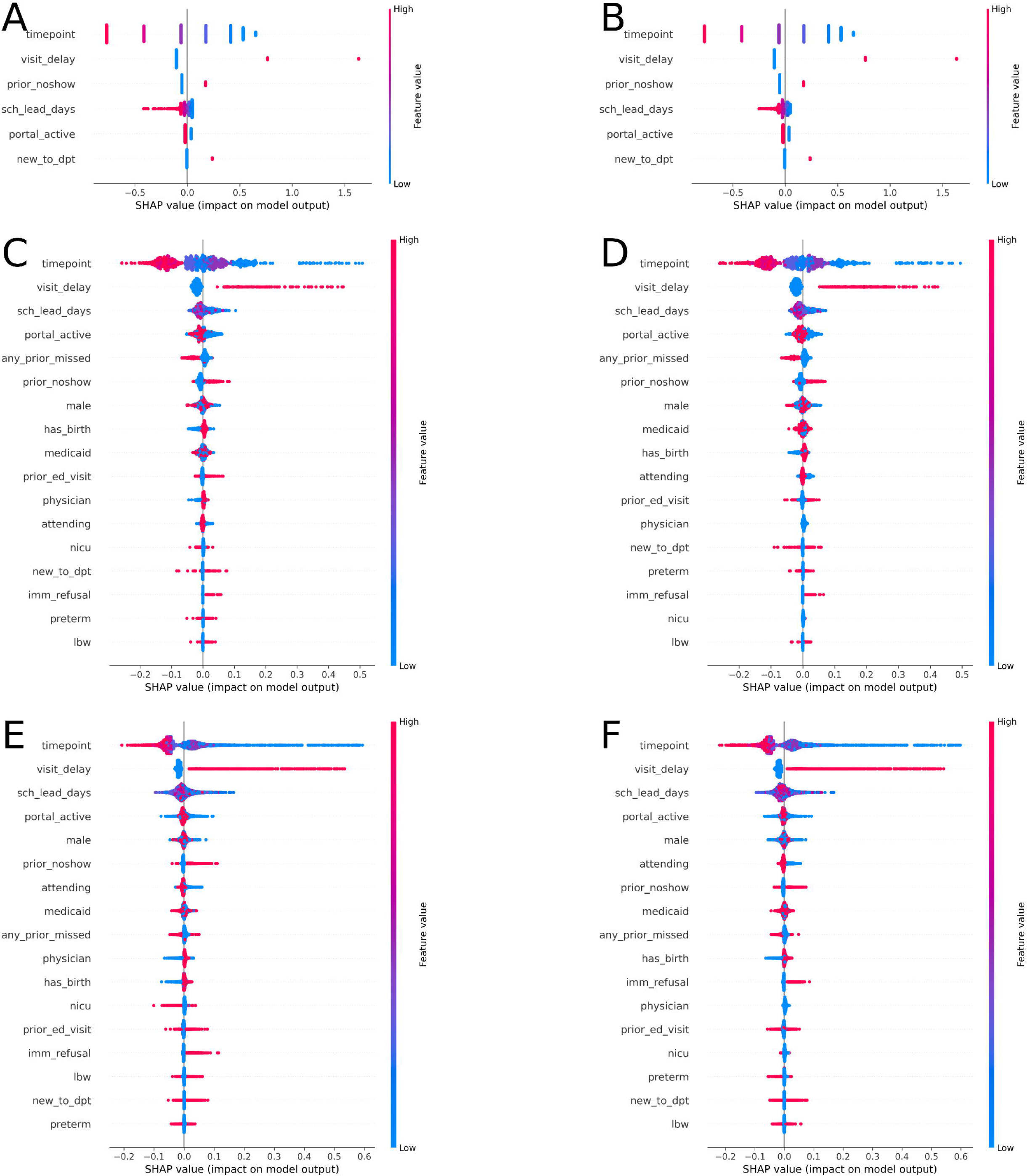
SHAP Bee Swarm Plots for Internal and External Validation Sets. **Legend:** SHAP values are shown for regularized logistic regression (A, B), random forest (C, D), and XGBoost (E, F). The left column presents internal validation using out of fold predictions from Practice A (A, C, E), and the right column presents external validation using predictions from the full Practice B cohort (B, D, F). Features are ordered by mean absolute SHAP value within each panel. Variable labels correspond to model covariates and include: timepoint (expected age in months at the current visit), visit_delay (months since the expected month of visit), prior_noshow (history of no show visits), sch_lead_days (days between scheduling and the current visit), new_to_dpt (new to the practice at the current appointment), imm_refusal (documented immunization refusal at the current or a prior visit), any_prior_missed (nonattendance at a previous well child visit), portal_active (patient portal activated), male (patient sex), medicaid (Medicaid insurance), prior_ed_visit (prior in system emergency department visit), attending (provider was an attending), physician (provider was a physician), has_birth (birth record in the health system), nicu (history of neonatal intensive care unit admission), preterm (gestational age <37 weeks), and lbw (birth weight <2,500 g).

Model discrimination was similar across insurance groups (AUC gap 0.004-0.024) and between English– and Spanish-speaking patients (**Table 3**). Among racial and ethnic groups, discrimination was comparable or modestly higher for Hispanic and non-Hispanic Black patients relative to non-Hispanic white patients across all three models. Point estimates were lower for Asian patients (AUC 0.49-0.57) and for patients whose preferred language was not English or Spanish (AUC 0.41-0.47); however, these subgroups contained fewer than 160 observations each, and the resulting confidence intervals were wide, overlapping substantially with those of larger subgroups. These estimates are therefore too imprecise to support conclusions about differential model performance.

**TABLE 3.** External Validation Model Discrimination by Patient Subgroup.

| Feature | Category | n | LR AUC | RF AUC | XGB AUC |
| --- | --- | --- | --- | --- | --- |
| Race/<br>Ethnicity | Hispanic | 1969 | 0.6636<br>(0.6332, 0.6941) | 0.6790<br>(0.6492, 0.7087) | 0.6749<br>(0.6463, 0.7036) |
|  | Black, non-Hispanic | 2390 | 0.6840<br>(0.6576, 0.7111) | 0.7008<br>(0.6754, 0.7275) | 0.7039<br>(0.6777, 0.7305) |
|  | White, non-Hispanic | 703 | 0.6140<br>(0.5618, 0.6594) | 0.6312<br>(0.5842, 0.6779) | 0.6273<br>(0.5792, 0.6743) |
|  | Asian, non-Hispanic | 77 | 0.5461<br>(0.3636, 0.7306) | 0.5748<br>(0.3898, 0.7588) | 0.4887<br>(0.3127, 0.6743) |
|  | Another Race,<br>non-Hispanic | 76 | 0.6301<br>(0.4423, 0.8108) | 0.6411<br>(0.4648, 0.7989) | 0.6191<br>(0.4387, 0.7811) |
|  | Not Reported,<br>non-Hispanic | 59 | 0.5362<br>(0.3714, 0.6937) | 0.6342<br>(0.4790, 0.7757) | 0.6408<br>(0.4792, 0.7885) |
| Preferred<br>Language | English | 4913 | 0.6712<br>(0.6534, 0.6895) | 0.6855<br>(0.6673, 0.7035) | 0.6837<br>(0.6650, 0.7020) |
|  | Spanish | 208 | 0.6453<br>(0.5553, 0.7353) | 0.6820<br>(0.5922, 0.7715) | 0.6963<br>(0.6045, 0.7804) |
|  | Another<br>Language | 153 | 0.4056<br>(0.3034, 0.5132) | 0.4726<br>(0.3587, 0.5867) | 0.4582<br>(0.3515, 0.5672) |
| Insurance | Not Medicaid | 1469 | 0.6789<br>(0.6406, 0.7156) | 0.6962<br>(0.6597, 0.7324) | 0.6791<br>(0.6414, 0.7145) |
|  | Medicaid | 3805 | 0.6551<br>(0.6337, 0.6762) | 0.6719<br>(0.6521, 0.6924) | 0.6749<br>(0.6536, 0.6956) |
| AUC Gap | Race/Ethnicity |  | 0.1478 | 0.126 | 0.2152 |
|  | Preferred<br>Language |  | 0.2656 | 0.2129 | 0.2381 |
|  | Insurance |  | 0.0238 | 0.0243 | 0.0042 |

**Note:** AUC, area under the receiver operating curve; LR, logistic regression (LASSO); RF, random forest; XGB, XGBoost. Metrics are reported for the external validation set. Confidence intervals were estimated using the percentile bootstrap (2,000 replicates). AUC Gap is defined as the difference between the highest and lowest subgroup AUC within each grouping variable.

## Discussion

We evaluated three models: regularized logistic regression, random forest, and XGBoost within this contextual framework integrating patient and clinical level data during an active appointment. We found that all models performed similarly, in both the internal and external validation sets. The model’s recall, or ability to identify positive cases, was modest; however the high negative predictive value provides a framework for sustainable resource allocation. In resource-constrained settings, identifying lower risk families allows health systems to safely de-prioritize intensive outreach for those likely to remain engaged in care. Further, while our reported metrics were selected to maximize F1 score, a value that optimizes precision and recall, clinics may choose to adjust their thresholds for greater specificity, which may be useful when outreach resources are limited.

Because these models predict nonattendance at the next recommended timepoint rather than no-show at a scheduled appointment, they identify risk while the family is still physically present and engaged in the medical home. This creates a window in which clinicians can address structural barriers to the next visit, such as coordinating transportation or scheduling a warm handoff to social work, before the family leaves the clinic. Children who are never captured by a scheduling system, and therefore invisible to no-show models, are included in this framing as long as they attended a prior visit. Additional applications for a model such as this include supporting transitions from inpatient or acute care settings to primary care by allowing hospitalists or emergency physicians to identify patients who are unlikely to attend a subsequent well-child visit and may benefit from additional supports, such as scheduling before discharge, reminders, or care coordination.

In infants, WCV attendance largely reflects parent or caregiver behavior, and the features that most influenced prediction were consistent with this. Scheduling patterns, visit delay, and prior no-shows indicate how families interact with the healthcare system, whereas infant clinical characteristics, including neonatal health status, were available to the models but contributed less. We did not include diagnostic or medication histories because these visits occurred during early infancy, when medical histories are limited and the primary reason for care is preventive. We also excluded area-level and social risk variables to avoid reintroducing demographic information that was intentionally omitted from the models.

Given that the outcome reflects family behavior rather than a clinical outcome, the observed AUC of 0.66-0.68 in external validation should be interpreted in the context of what is achievable when predicting behavioral outcomes. Models predicting disease from clinical biomarkers or physiological signals routinely achieve AUCs above 0.80, but discrimination declines substantially when the outcome depends on human behavior and contextual decision-making rather than pathophysiology.^31–34^ Across clinical domains, models predicting patient behaviors such as treatment response, adherence, and substance use in external validation settings have reported AUCs in the range of 0.64-0.74, even when using richer feature sets or more complex modeling approaches.^35–37^ The discrimination observed in this study is consistent with these expectations and reflects the inherent difficulty of predicting behavioral outcomes from routine clinical data.

A common issue in implementing risk stratification or clinical decision support tool is integration in clinical workflows and in currently-used technology.^38,39^ Electronic health records are often limited in the types of models that are supported. For instance, many EHR systems allow implementation of customer-created models, but use of complex machine learning models such as tree-based models often require additional licenses to support the increased computational needs. Simpler models such as logistic regression are often simpler to implement; therefore, it is attractive from an implementation perspective if the simpler logistic regression model has similar predictive performance.^40^

Similarly, reducing clinician burden by requiring fewer measures to be input into a model may promote uptake of risk stratification tools.^41^ Our logistic regression model with LASSO regularization ultimately contained six variables that meaningfully contributed to prediction (i.e. nonzero coefficients): timepoint, time from when the current visit was due and the date, prior no-shows, how many days in advance the visit was scheduled, whether the patient is new to the department, and whether the patient has a documented immunization refusal. These primarily comprise care delivery features which are easily accessible to the clinician and would not require significant time to identify. If embedded in the EHR, it is also possible to have these measures automatically filled, to further facilitate use without increasing clinician burden. A prediction model could also be used in concert with other reminder systems that are already common strategies used to improve visit attendance among health systems.^42^

Model discrimination was similar across most subgroups, including insurance type and the two largest racial/ethnic groups in the validation cohort. Point estimates of AUC were lower for Asian patients and those with non-English/non-Spanish language preference, but confidence intervals for these subgroups were wide and overlapped with those of larger groups, preventing conclusions about differential performance. If these differences are real, plausible explanations include the heterogeneity of the Asian racial category, which groups together families from many different countries, languages, and cultures, and the possibility that care delivery features like scheduling patterns and portal activation function differently when families navigate the system through an interpreter. Confirming whether model performance differs for these populations will require validation in cohorts with larger subgroup representation.

A limitation of this research is that the external validation set arises from the same vertically integrated healthcare system. While clinics may have different patient populations, providers, and internal practices, there may be system-level factors, such as the way measures are stored in the EHR, that make the two clinics more similar than among those in different healthcare systems. We did not evaluate drift in our predictions. There may be changes in how people use healthcare over time, meaning that our model trained on births from 2022 may not continue to be accurate over time. Future work should explore whether these models maintain performance when tested on cohorts from different period. Additionally, the training cohort underrepresented certain subgroups, limiting predictive performance. Future validation should include training data with broader demographic and linguistic representation and evaluate performance against geographically and institutionally independent cohorts.

Predicting WCV nonattendance at the next recommended timepoint is feasible using routine visit data, and a parsimonious logistic regression model performs comparably to more complex algorithms. The most influential predictors across all models were care delivery features available at the point of care, which may facilitate EHR integration without increasing clinician burden. Unlike no-show models, this approach identifies risk while the family is still present in the medical home, creating an opportunity for clinicians to address barriers to the next visit before a lapse in preventive care occurs.

## Author Contributions

**Amanda Luff:** Conceptualization, Methodology, Data Curation, Formal analysis, Writing – Original Draft, Visualization. **Maureen Shields:** Writing – Original Draft. **Jana Hirschtick:** Writing – Original Draft. **Marybeth Ingle:** Funding acquisition, Writing – Original Draft. **Clare Crosh:** Funding acquisition, Writing – Original Draft. **Melanie Marsh:** Writing – Original Draft. **Francois Modave:** Methodology, Writing – Original Draft. **Veronica Fitzpatrick:** Funding acquisition, Supervision, Writing – Original Draft.

## Data Availability

The data underlying this study contain protected health information collected during routine clinical care and cannot be shared publicly due to HIPAA restrictions. Requests for de-identified data may be directed to the corresponding author and would be subject to institutional review, including data use agreements and regulatory approvals. Analytic code is available at https://doi.org/10.5281/zenodo.20648820.43

## Supplementary Tables

**S1 Table.** Hyperparameter Tuning Methods and Best Estimator Specifications

**S2 Table.** Threshold-Specific Metrics at a 0.5 Threshold

## Notes

### Competing Interest Statement

The authors have declared no competing interest.

### Summary of Updates

Addition of subgroup performance metrics

