## Supplementary material for "Predicting Infant Nonattendance at the Next Recommended Well-Child Visit: Model Development and Validation": Supp Table 1

**Supplementary Table 1.** Hyperparameter Tuning Methods and Best Estimator Specifications

| Model | Grid Search Method | Ideal Hyperparameters |
|---|---|---|
| Logistic Regression | Full Grid Search<br>penalty: L1, L2<br>C: 0.001, 0.01, 0.02, 0.05, 0.1, 0.5, 1, 3, 10 | <br>penalty: L1<br>C: 0.01 |
| Random Forest | Random Grid Search (300 iterations)<br>max_depth: 10, 20, 50, 70, 80, 100, 150<br>max_features: 2, 4, 6, sqrt, log2<br>min_samples_leaf: 2, 5, 7, 10, 15<br>min_samples_split: 2, 5, 7, 10, 12, 15, 18, 20<br>n_estimators: 100, 200, 500, 1000, 1500, 2000 | <br>max_depth: 10<br>max_features: 6<br>min_samples_leaf: 2<br>min_samples_split: 5<br>n_estimators: 2000 |
| XGBoost | Random Grid Search (300 iterations)<br>max_depth: 3, 5, 7, 9, 11<br>learning_rate: 0.05, 0.1, 0.2<br>n_estimators: 100, 300, 500, 800, 1000, 1200<br>subsample: 0.4, 0.6, 0.8<br>colsample_bytree: 0.4, 0.5, 0.6<br>min_child_weight: 2, 3, 5, 8<br>gamma: 0, 1, 2, 3, 4, 5<br>reg_alpha: 0, 0.5, 1, 2, 3<br>reg_lambda: 0, 0.5, 1, 2, 3<br>scale_pos_weight: 0.5, 1, 2, 5 | <br>max_depth: 11<br>learning_rate: 0.1<br>n_estimators: 300<br>subsample: 0.8<br>colsample_bytree: 0.4<br>min_child_weight: 2<br>gamma: 0<br>reg_alpha: 2<br>reg_lambda: 2<br>scale_pos_weight: 0.5 |

*All searches used nested group k-fold cross validation with grouping at the patient level and k=5. Scoring was based on average precision. Continuous variables were standardized using StandardScaler for Logistic Regression. Class weight was specified as balanced for Logistic Regression and Random Forests. SMOTE was used to improve class balance for Random Forests and XGBoost.*
