## Supplementary material for "Predicting Infant Nonattendance at the Next Recommended Well-Child Visit: Model Development and Validation": Supp Table 2

**S2 Table.** Threshold-Specific Metrics at a 0.5 Threshold

| Model | Accuracy | Balanced Accuracy | Sensitivity (Recall) | Specificity | PPV (Precision) | NPV | F1 |
|---|---|---|---|---|---|---|---|
| **Internal Validation (Practice A, Out-of-Fold)** | | | | | | | |
| **LR** | 0.5568 (0.5423, 0.5696) | 0.6128 (0.5951, 0.6288) | 0.6957 (0.6631, 0.7255) | 0.5298 (0.5145, 0.5444) | 0.2230 (0.2070, 0.2395) | 0.8998 (0.8879, 0.9105) | 0.3378 (0.3174, 0.3585) |
| **XGB** | 0.8511 (0.8411, 0.8609) | 0.5638 (0.5522, 0.5752) | 0.1382 (0.1153, 0.1605) | 0.9894 (0.9862, 0.9923) | 0.7169 (0.6455, 0.7833) | 0.8555 (0.8454, 0.8653) | 0.2317 (0.1971, 0.2646) |
| **RF** | 0.8445 (0.8349, 0.8540) | 0.6123 (0.5976, 0.6276) | 0.2683 (0.2392, 0.2977) | 0.9563 (0.9503, 0.9621) | 0.5435 (0.4966, 0.5918) | 0.8708 (0.8607, 0.8804) | 0.3593 (0.3259, 0.3928) |
| **External Validation (Practice B)** | | | | | | | |
| **LR** | 0.5612 (0.5480, 0.5738) | 0.6068 (0.5909, 0.6220) | 0.6846 (0.6568, 0.7115) | 0.5289 (0.5136, 0.5436) | 0.2756 (0.2584, 0.2924) | 0.8650 (0.8519, 0.8772) | 0.3930 (0.3726, 0.4127) |
| **XGB** | 0.8017 (0.7909, 0.8123) | 0.5486 (0.5398, 0.5587) | 0.1161 (0.0987, 0.1354) | 0.9811 (0.9769, 0.9855) | 0.6165 (0.5539, 0.6859) | 0.8092 (0.7985, 0.8198) | 0.1954 (0.1689, 0.2247) |
| **RF** | 0.7857 (0.7744, 0.7965) | 0.5784 (0.5657, 0.5908) | 0.2239 (0.2002, 0.2475) | 0.9328 (0.9251, 0.9406) | 0.4658 (0.4244, 0.5069) | 0.8212 (0.8097, 0.8319) | 0.3025 (0.2741, 0.3300) |

**Note:** PPV, Positive Predictive Value; NPV, Negative Predictive Value; LR, logistic regression; RF, random forest; XGB, XGBoost. Confidence intervals were estimated using the percentile bootstrap (2,000 replicates).
